# Ensitrelvir Fumaric Acid in Patients with SARS-CoV-2: A Retrospective Chart Review

**DOI:** 10.1101/2023.09.03.23294865

**Authors:** Masaya Yamato, Masahiro Kinoshita, Shogo Miyazawa, Masayuki Seki, Tomoki Mizuno, Takuhiro Sonoyama

## Abstract

**Introduction:** Antivirals with differing mechanisms of action are needed for COVID-19 treatment; remdesivir is mainly used in hospitalized patients, but additional antivirals are needed in this setting. Ensitrelvir, a 3C-like protease inhibitor, received emergency approval in Japan in November 2022, based on evidence of rapid symptom resolution in non-hospitalized patients, but confirmation of its efficacy in hospitalized patients is lacking.

**Case presentations:** This case series reports outcomes for all patients who received ensitrelvir whilst hospitalized with SARS-CoV-2 infection at Rinku General Medical Center, Japan (November 2022– April 2023). Thirty-two hospitalized patients received five days of ensitrelvir treatment (375/125 mg). Mean age was 73.5 years and most patients had mild COVID-19. Patients exhibited various underlying diseases, most commonly hypertension (78.1%) and chronic kidney disease (25.0%). Seven (21.9%) patients were on hemodialysis. The most common concomitant medications were antihypertensives (59.4%) and corticosteroids (31.2%); two (6.3%) patients were being treated with rituximab; 27 (84.4%) patients had viral persistence following pre-treatment remdesivir failure. Following ensitrelvir treatment, all patients experienced clinical improvement as assessed by the investigator. No ICU admissions or deaths due to COVID-19 occurred. Viral clearance was recorded in 18 (56.3%) patients by day 5 and 25 (78.1%) patients at final measurement. No new safety signals were observed.

**Discussion:** This case series represents ensitrelvir clinical efficacy in hospitalized patients with SARS-CoV-2 in a real-world setting. Most patients experienced persistent viral shedding despite pre-treatment with antivirals, and most were considered high-risk due to underlying conditions. Despite this, no patients experienced progression of COVID-19 to severe or critical disease, including those who failed prior remdesivir treatment. The antiviral activity of ensitrelvir demonstrated here indicates it is a viable treatment option for SARS-CoV-2 infection in this setting.

**Conclusion:** Ensitrelvir was associated with potent antiviral activity and positive clinical outcomes in high-risk, hospitalized patients with various comorbidities.

**Trial Registration:** UMIN000051300

**Key Summary Points:** *Why carry out this study?:*

- Real-world evidence of ensitrelvir for the treatment of COVID-19, especially in inpatient settings, is required.
- This was a retrospective cohort analysis, consisting of 32 high-risk hospitalized patients with a range of disease severities and comorbidities, including patients who had experienced remdesivir treatment failure.

*What was learned from the study?:*

- All patients experienced clinical improvement following ensitrelvir treatment, without any worsening COVID-19.
- Viral clearance was documented in 78.1% of patients at final viral measurement.
- No new safety signals of ensitrelvir were observed in this analysis.

## INTRODUCTION

The first cases of the novel coronavirus disease (COVID-19) caused by severe acute respiratory syndrome coronavirus 2 (SARS-CoV-2) infection were identified in Wuhan City, Hubei Province, China, in December 2019[1]. Following the global spread of infections, the World Health Organization (WHO) declared COVID-19 a pandemic in March 2020[2]. As of June 2023, there have been ∼768,000,000 confirmed cases and ∼6,900,000 deaths reported to the WHO[3], including ∼33,800,000 cases and ∼75,000 deaths reported in Japan[3]. Since its identification as a variant of concern (VOC) in November 2021, the Omicron variant and its sublineages have been the dominant SARS-CoV-2 variants worldwide, responsible for the vast majority of reported cases[4–6]. Compared with previous VOC’s (particularly Delta), the Omicron variant is associated with reduced symptom severity[7–10], but increased transmission[11–13] and immune evasion[14, 15]. Whilst Omicron infection generally leads to less severe outcomes than Delta, its increased transmissibility led to a significant healthcare burden globally; Japan’s sixth, seventh, and eighth COVID-19 waves, caused by Omicron infection, were associated with the highest levels of COVID-19 cases and deaths reported since the start of the pandemic[3].

Booster vaccines and hybrid immunity (immunity developed from both vaccination and previous SARS-CoV-2 infection) consistently provide high protection against Omicron-related severe disease, but their effectiveness wanes over time[16–18]; furthermore, many monoclonal antibodies previously used to treat people infected with SARS-CoV-2 have demonstrated reduced neutralization and binding affinity to the Omicron variant and its sublineages[19, 20]. Patients with severe comorbidities are still at increased risk of poor outcomes when infected with the Omicron variant, including progression to severe disease and death[21–23]. Additionally, patients with severe comorbidity burden and immunocompromised patients, including those on hemodialysis[24], are likely to be at more risk from prolonged viral shedding and viral persistence[25–29]. This can increase the overall disease burden for these patients, for example by interfering with the management of underlying diseases in patients with hematological malignancies[30, 31]. An antiviral agent that can rapidly reduce SARS-CoV-2 virus levels and prevent viral persistence would therefore be a beneficial treatment option for this patient population.

Remdesivir is an intravenously administered antiviral, which is indicated in Japan for the treatment of COVID-19 in patients: with oxygen saturation ≤ 94%; who require supplementary oxygen; or with pneumonia because of SARS-CoV-2[32]. Nirmatrelvir/ritonavir (Paxlovid®) and molnupiravir (Lagevrio®) are oral antivirals for the treatment of COVID-19; both have received emergency authorization in many countries, including Japan, for the treatment of patients infected with SARS-CoV-2, with risk factors for severe disease. Nirmatrelvir/ritonavir also received the United States Food and Drug Administration’s full approval in May 2023[33]. In clinical trials in non-hospitalized patients, both drugs demonstrated significantly lower rates of hospitalization or death versus placebo[34, 35], and reductions in SARS-CoV-2 viral ribose nucleic acid by day 5[34, 36]. However, approval for both drugs was based on clinical trials in unvaccinated individuals, before Omicron became the predominant SARS-CoV-2 strain[34, 35]. Since then, nirmatrelvir/ritonavir failed to meet the primary endpoint of self-reported, sustained alleviation of all symptoms in non-hospitalized patients at low risk of progressing to severe disease in the phase 2/3 EPIC-SR study[37].

Ensitrelvir fumaric acid (Xocova®) is an oral antiviral, developed by Shionogi & Co., Ltd. for the treatment of infection caused by SARS-CoV-2. Ensitrelvir acts through the same mechanism as nirmatrelvir, inhibiting the 3C-like protease, which is essential for viral replication and processing of the polyprotein encoded by the SARS-CoV-2 gene[38]. In the recently reported phase 3 trial SCORPIO-SR, ensitrelvir met the primary endpoint and demonstrated a significantly shorter time to resolution of five symptoms typical of Omicron infection compared with placebo (−24.3 hours; 95% CI − 78.7 to 11.7; *p* = 0.04) in patients aged 12–69 with mild-to-moderate COVID-19[39]. Based on these data, as well as efficacy from the phase 2b part[40], ensitrelvir received emergency authorization in Japan for the treatment of SARS-CoV-2 infection. In addition, ensitrelvir has demonstrated rapid and significant reductions in SARS-CoV-2 viral RNA: a 30-fold reduction in mean viral RNA levels compared with placebo at day 4 [mean difference –1.47 log_10_ tissue culture infectious dose (TCID_50_)/mL] was recorded in the SCORPIO-SR trial[39]. Ensitrelvir treatment in hospitalized patients is being investigated in the ongoing STRIVE study[41].

Despite positive outcomes reported in clinical trials, evidence on the efficacy and safety of ensitrelvir in clinical practice is limited. The aim of this retrospective chart review was to evaluate the treatment status, including patient background and treatment outcomes, of patients treated with ensitrelvir in Japan, November 2022 and April 2023, immediately after emergency approval of ensitrelvir in November 2022 and when the Omicron variant was dominant.

## METHODS

### Study Design/Period

This was a single center, retrospective chart review of patients prescribed ensitrelvir for the treatment of SARS-CoV-2 infection, from November 2022 to April 2023 in the Rinku General Medical Center. Ensitrelvir (375/125 mg) was dosed orally once a day for five days, where 375 mg was dosed on day 0 as a loading dose and 125 mg was dosed on days 1−4 as a maintenance dose.

### Patient Population

All patients who received ensitrelvir between November 2022 and April 2023 were included in the study. Only patients for whom the following data were obtained were included in the analytical population (Fig. 1): start and end date of ensitrelvir treatment; a SARS-CoV-2 positive test result; SARS-CoV-2 virus test results [antigen test, reverse transcription polymerase chain reaction (RT-PCR)] after starting administration of ensitrelvir; clinical outcomes of SARS-CoV-2 infection. Patients were excluded from the analytical population if they were not treated in accordance with ensitrelvir prescribing instruction or if they refused to participate in the study. This current paper reports results for hospitalized patients only.

**Fig. 1.**
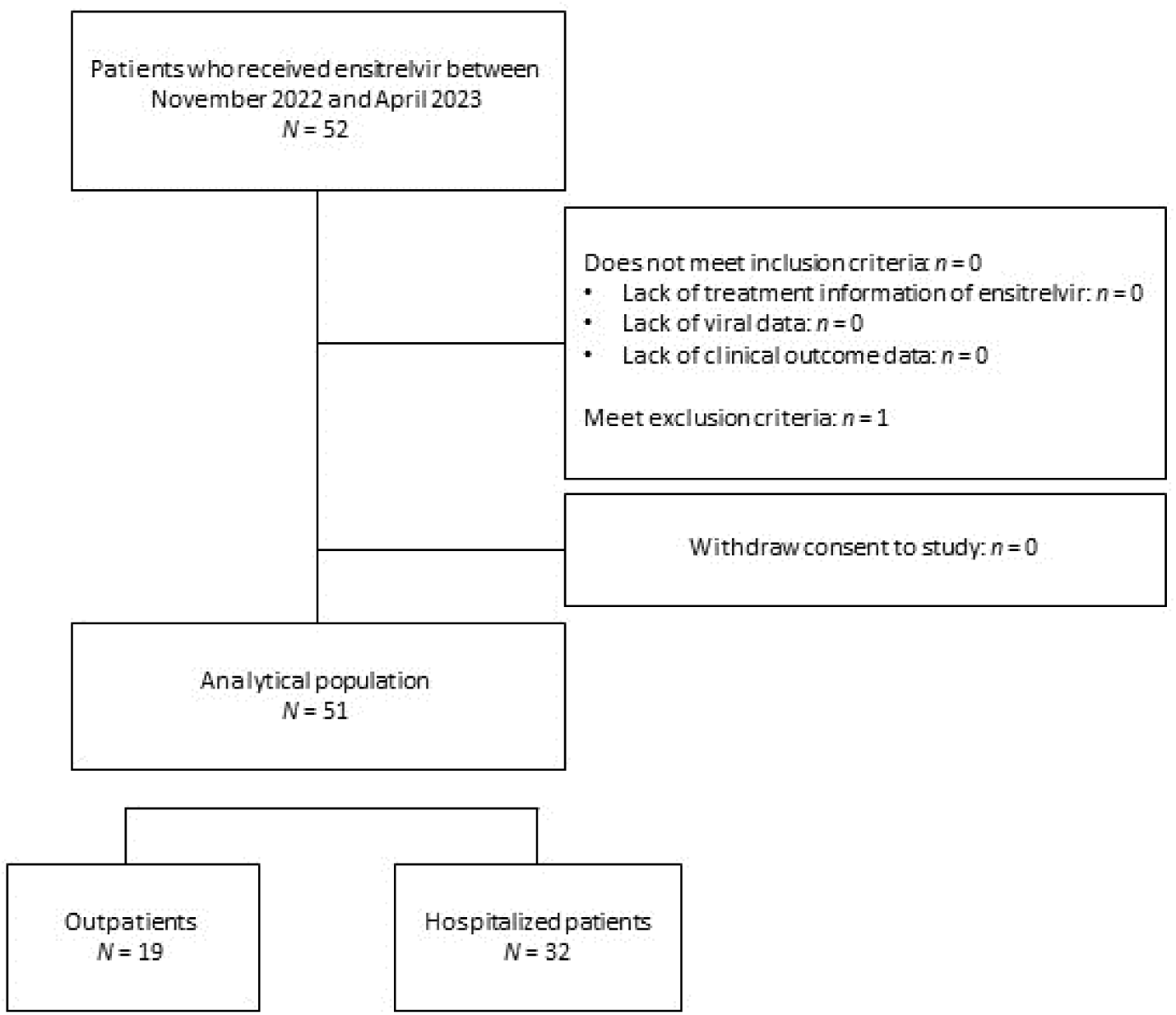
Patient disposition flowchart.

### Data Collection

Data were collected using electronic medical records of the Rinku General Medical Center. Pseudonymized information was aggregated and analyzed as anonymized information using an anonymization tool (CoNaxs®, Conax Automomation Sdn. Bhd).

The study collected general information of patients including gender, age, date of COVID-19 onset, underlying disease/complications, pre-treatment and post-treatment for SARS-CoV-2 infection, concomitant medications, previous SARS-CoV-2 infection history, and SARS-CoV-2 vaccination status (number of vaccinations and time of last vaccination).

Treatment information collected included ensitrelvir administration start and end date, prior therapies, and concomitant drug use.

Clinical information was also collected, including imaging findings, body temperature (axillary), clinical signs and symptoms, level of consciousness, and oxygen saturation. Disease severity at the start of ensitrelvir administration was classified by investigator according to the Japanese guideline on COVID-19 treatment (Supplementary Table 1)[42]. The presence/absence of pneumonia, oxygen administration (including flow rate), mechanical ventilation, and intensive care unit (ICU) management were also recorded, as well as the implementation status of dialysis. Finally, clinical outcomes of SARS-CoV-2 infection and any adverse events (AEs) related to ensitrelvir were recorded.

Virological information collected consisted of SARS-CoV-2 virus test results (antigen test, RT-PCR). Quantitative antigen level in nasopharyngeal swab was assessed using Lumipulse^®^ (Fujirevio, Tokyo, Japan), with a maximum viral antigen level of 5,000 pg/mL and a cut-off value of 1.34 pg/mL. It has previously been reported that a cut-off value of 89.73 pg/mL corresponds to a cycle threshold (Ct) value of ≥ 35 by quantitative PCR[43]. When estimating the infectivity of SARS-CoV-2, a Ct value ≥ 35 is often used as a marker for non-infectious viral levels[44–46]. Therefore, we defined viral clearance as an antigen level of < 89.73 pg/mL. Viral rebound was defined as an increase of antigen level of ≧ 89.73 pg/mL after viral clearance.

### Endpoints

The primary endpoint of the study was viral antigen or PCR results (quantitative or qualitative results) at each time point from the start of administration to 10 days after the end of ensitrelvir administration. The secondary endpoint was investigator-assessed improvement at the end of ensitrelvir treatment (day 4 following administration of ensitrelvir). Exploratory endpoints included presence or absence of hospitalization; ICU admission; and death up to 14 days after the start of administration. Safety of ensitrelvir treatment was also assessed, by recording any treatment-related AEs. No statistical analyses were performed as this was a descriptive study.

## RESULTS

### Patient disposition and baseline characteristics

Fifty-two patients with SARS-CoV-2 infection who received ensitrelvir were identified between November 2022 and April 2023 at Rinku General Medical Center. One was excluded due to co-usage of contraindicated drugs. In total, 32 of the included patients were hospitalized and 19 were outpatients (Fig. 1). Here we report the outcomes for the 32 hospitalized patients only, all of whom completed 5 days of ensitrelvir treatment. Outcomes for the 19 outpatients will be reported in detail separately, due to the limited follow-up data available.

Within the hospitalized analytical population (*N* = 32), the average age was 73.5 years old, and 25 (78.1%) patients were age 65 years and older (Table 1). At the start of ensitrelvir treatment, 21/32 (65.6%) patients had mild COVID-19 disease severity; four (12.5%) had moderate I severity; and seven (21.9%) had moderate II severity (Table 1), according to the Japanese guideline. All seven patients with moderate II disease were on inhaled oxygen therapy and eight patients (25%) had evidence of pneumonia at the start of ensitrelvir treatment (two with moderate I disease and six with moderate II) (Table 1). Prior antiviral therapy had been administered to 29/32 (90.6%) patients, including all 11 patients with moderate I or II disease (Table 1). The most common prior therapy was remdesivir [27/32 (84.4%)], either delivered as monotherapy [*n* = 24 (75%)] or alongside casirivimab/imdevimab [*n* = 3 (9.4%)], and 21/27 (77.8%) patients received ≧ 3 days of remdesivir (mean 6.6 days, range 3–22 days). One patient (3.1%) received casirivimab/imdevimab monotherapy and one patient (3.1%) received molnupiravir monotherapy. Patients were switched to ensitrelvir based on investigator assessments of clinical and virological status: either clinical symptoms were not resolved, or viral clearance was required for further medical procedures.

**Table 1.**
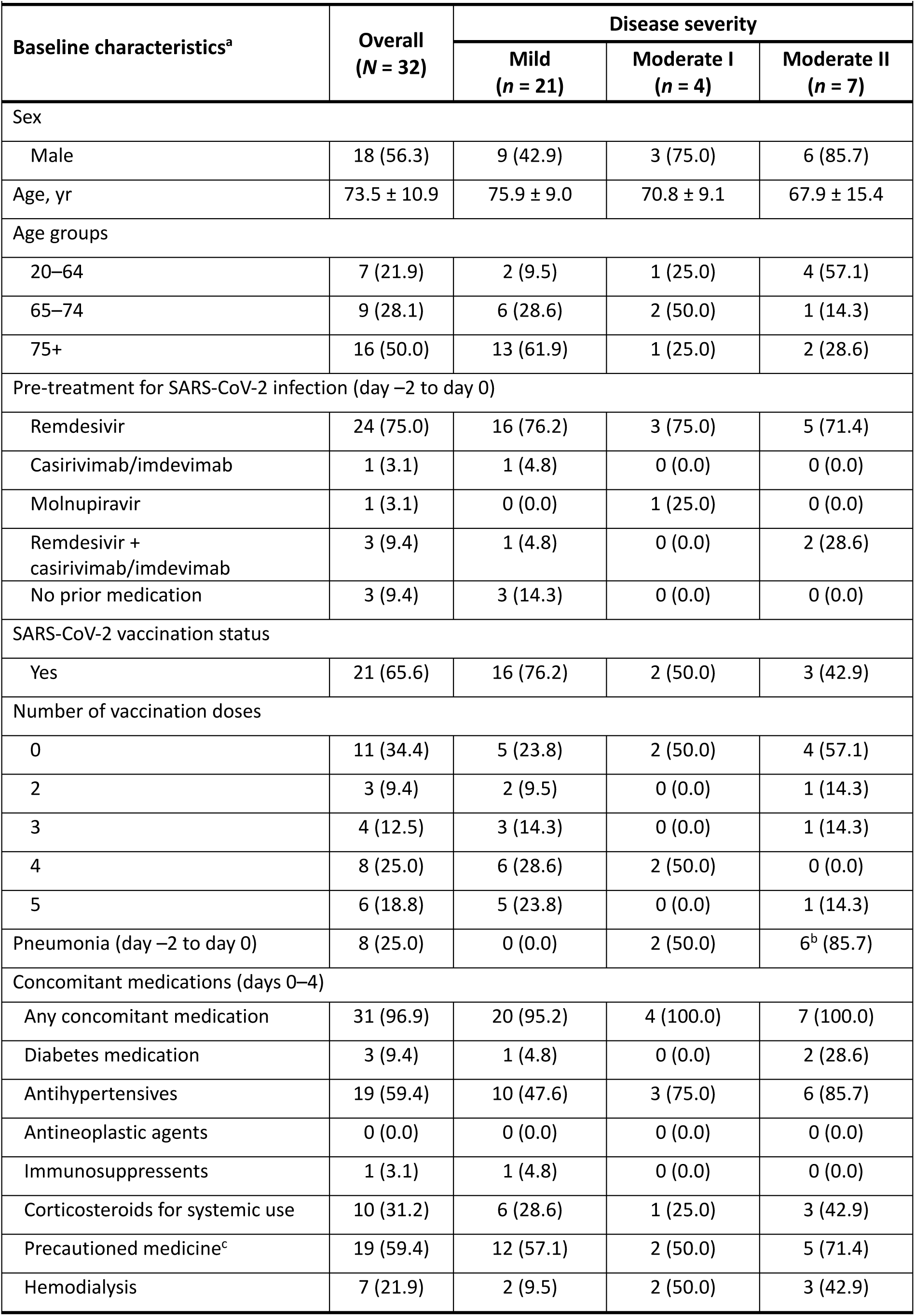
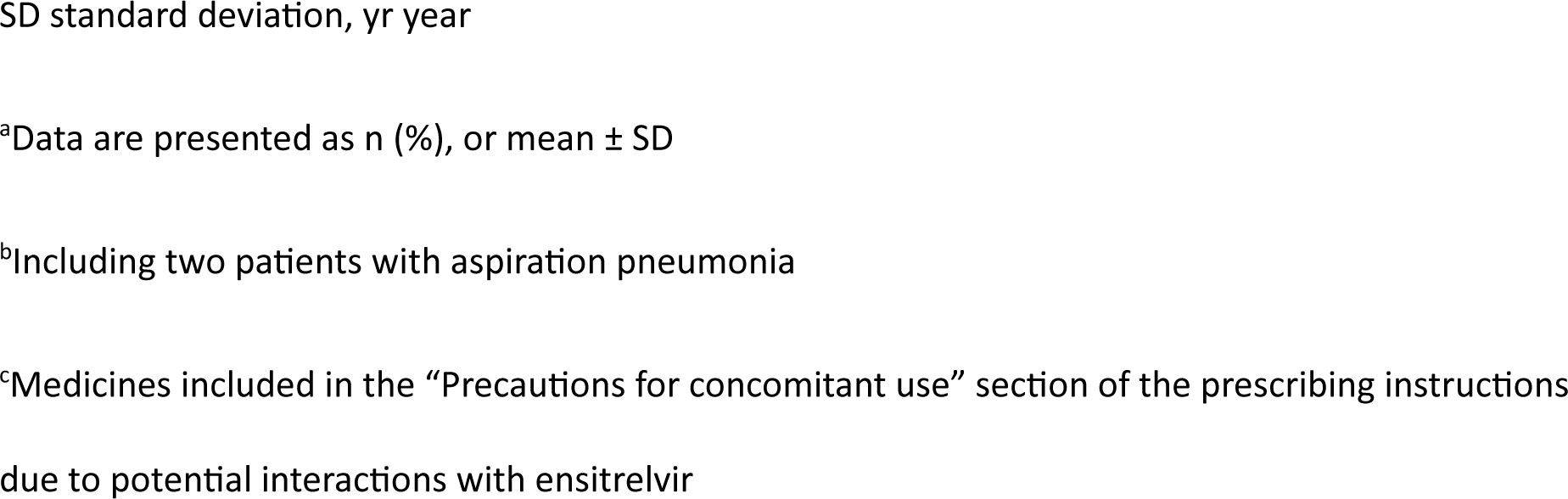
Demographics and baseline characteristics overall, and by disease severity at start of ensitrelvir treatment.

| Baseline characteristics<sup>a</sup> | Overall<br>(N = 32) | Disease severity |  |  |
| --- | --- | --- | --- | --- |
|  |  | Mild<br>(n = 21) | Moderate I<br>(n = 4) | Moderate II<br>(n = 7) |
| Sex |  |  |  |  |
| Male | 18 (56.3) | 9 (42.9) | 3 (75.0) | 6 (85.7) |
| Age, yr | 73.5 ± 10.9 | 75.9 ± 9.0 | 70.8 ± 9.1 | 67.9 ± 15.4 |
| Age groups |  |  |  |  |
| 20–64 | 7 (21.9) | 2 (9.5) | 1 (25.0) | 4 (57.1) |
| 65–74 | 9 (28.1) | 6 (28.6) | 2 (50.0) | 1 (14.3) |
| 75+ | 16 (50.0) | 13 (61.9) | 1 (25.0) | 2 (28.6) |
| Pre-treatment for SARS-CoV-2 infection (day –2 to day 0) |  |  |  |  |
| Remdesivir | 24 (75.0) | 16 (76.2) | 3 (75.0) | 5 (71.4) |
| Casirivimab/imdevimab | 1 (3.1) | 1 (4.8) | 0 (0.0) | 0 (0.0) |
| Molnupiravir | 1 (3.1) | 0 (0.0) | 1 (25.0) | 0 (0.0) |
| Remdesivir +<br>casirivimab/imdevimab | 3 (9.4) | 1 (4.8) | 0 (0.0) | 2 (28.6) |
| No prior medication | 3 (9.4) | 3 (14.3) | 0 (0.0) | 0 (0.0) |
| SARS-CoV-2 vaccination status |  |  |  |  |
| Yes | 21 (65.6) | 16 (76.2) | 2 (50.0) | 3 (42.9) |
| Number of vaccination doses |  |  |  |  |
| 0 | 11 (34.4) | 5 (23.8) | 2 (50.0) | 4 (57.1) |
| 2 | 3 (9.4) | 2 (9.5) | 0 (0.0) | 1 (14.3) |
| 3 | 4 (12.5) | 3 (14.3) | 0 (0.0) | 1 (14.3) |
| 4 | 8 (25.0) | 6 (28.6) | 2 (50.0) | 0 (0.0) |
| 5 | 6 (18.8) | 5 (23.8) | 0 (0.0) | 1 (14.3) |
| Pneumonia (day –2 to day 0) | 8 (25.0) | 0 (0.0) | 2 (50.0) | 6<sup>b</sup> (85.7) |
| Concomitant medications (days 0–4) |  |  |  |  |
| Any concomitant medication | 31 (96.9) | 20 (95.2) | 4 (100.0) | 7 (100.0) |
| Diabetes medication | 3 (9.4) | 1 (4.8) | 0 (0.0) | 2 (28.6) |
| Antihypertensives | 19 (59.4) | 10 (47.6) | 3 (75.0) | 6 (85.7) |
| Antineoplastic agents | 0 (0.0) | 0 (0.0) | 0 (0.0) | 0 (0.0) |
| Immunosuppressents | 1 (3.1) | 1 (4.8) | 0 (0.0) | 0 (0.0) |
| Corticosteroids for systemic use | 10 (31.2) | 6 (28.6) | 1 (25.0) | 3 (42.9) |
| Precautioned medicine<sup>c</sup> | 19 (59.4) | 12 (57.1) | 2 (50.0) | 5 (71.4) |
| Hemodialysis | 7 (21.9) | 2 (9.5) | 2 (50.0) | 3 (42.9) |

SD standard deviation, yr year
<sup>a</sup>Data are presented as n (%), or mean ± SD
<sup>b</sup>Including two patients with aspiration pneumonia

A higher proportion of patients with moderate I or II disease [2/4 (50.0%) and 4/7 (57.1%), respectively] were unvaccinated for SARS-CoV-2, compared with patients with mild disease [5/21 (23.8%)] (Table 1). The most common concomitant medications were antihypertensives [19 (59.4%)] and corticosteroids [10 (31.2%)]. One (3.1%) patient received immunosuppressants (cyclosporine), and 18 (58.1%) received precautioned medicines (medicines included in the “Precautions for concomitant use” section of the prescribing instructions due to potential interactions with ensitrelvir). Two (6.3%) patients received rituximab in the days before ensitrelvir treatment (days –49 and –16, respectively). Seven (22.6%) patients received hemodialysis. Patients exhibited a wide range of underlying diseases: the most common underlying comorbidity was hypertension [25/32 (78.1%)] followed by chronic kidney disease [8/32 (25.0%)], cerebral infarction [7/32 (21.9%)], diabetes mellites [7/32 (21.9%)], and dyslipidemia [7 (21.9%)]. Ten patients (31.3%) had malignant tumor. Individual patient demographics and characteristics can be seen in Table 2.

### Virology outcomes

All 32 patients received the full five days of ensitrelvir treatment (days 0–4) (Table 3). Viral clearance (antigen level < 89.73 pg/mL) was observed by day 5 in 18/32 (56.3%) patients and antigen level below lower the limit of quantification (cut-off level: < 1.34 pg/mL) was observed in 5/32 (15.6%) patients by day 5. Viral clearance was observed at final measurement in 25/32 (78.1%) patients and a final antigen quantification measurement of < 1.34 pg/mL was observed in 20/32 (62.5%) patients.

**Table 3.** Virology outcomes.

| Patient | Quantitative viral antigen (pg/mL) |  |  |  |  |  |  |  |  |  |  |  |  |  |  |
| --- | --- | --- | --- | --- | --- | --- | --- | --- | --- | --- | --- | --- | --- | --- | --- |
|  | Days from start of ensitrelvir administration |  |  |  |  |  |  |  |  |  |  |  |  |  |  |
|  | Day -1<br>and<br>Day -2 | Day 0 | Day 1 | Day 2 | Day 3 | Day 4<br>(EOT) | Day 5 | Day 6 | Day 7 | Day 8 | Day 9 | Day 10 | Day 11 | Day 12 | Day 13 |
| 1 | 5000 | 5000 |  | 571.44 |  |  | 21.75 | 18.67 | 9.43 | 2.81 | 0.06 | 4.79 | 0.01 | 0.10 |  |
| 2 | 206.25 | 3429.86 | 235.08 | 10.35 | 1.06 | 0.95 |  |  |  |  |  |  |  |  |  |
| 3 | 5000 |  | 5000 |  |  | 113.88 |  | 0.01 |  |  |  |  |  |  |  |
| 4 | 255.09 | 5000 |  |  | 36.55 |  | 25.72 | 1.61 | 0.30 | 0.01 |  |  |  |  |  |
| 5 | 5000 | 5000 |  | 5000 | 5000 | 5000 |  |  |  | 1233.07 | 494.52 |  | 254.76 | 8.67 | 44.64 |
| 6 | 5000 | 5000 |  | 1394.43 |  | 39.12 | 43.49 | 46.30 |  | 28.65 | 0.83 | 0.89 |  |  |  |
| 7 |  | 5000 |  | 1676.19 |  |  |  |  | 27.73 |  |  |  |  |  |  |
| 8 | 5000 | 2728.74 | 3736.13 | 142.46 |  | 13.50 | 0.30 |  |  |  |  |  |  |  |  |
| 9 | 5000 | 5000 |  | 2587.88 |  | 4.77 | 1210.73 |  | 5000 | 5000 | 5000 |  |  |  |  |
| 10 |  | 1227.48 |  | 182.59 | 200.71 |  |  |  |  |  |  |  |  |  |  |
| 11 | 5000 | 5000 | 5000 |  | 1086.83 | 4091.46 |  |  | 16.27 | 11.83 | 3.76 | 1.73 | 0.01 |  |  |
| 12 | 5000 |  |  | 1.04 | 0.11 |  |  |  |  |  |  |  |  |  |  |
| 13 | 5000 |  | 5000 |  |  | 0.75 |  |  |  |  |  |  |  |  |  |
| 14 | 5000 | 5000 |  |  | 87.07 | 25.39 | 8.50 | 6.81 | 7.94 | 94.59 | 0.14 | 2.29 | 0.05 | 1.73 | 0.57 |
| 15 |  | 5000 |  |  | 5000 |  | 665.36 |  | 352.02 |  |  | 8.56 | 1.30 | 4.21 | 0.01 |
| 16 | 5000 | 5000 |  | 53.22 |  |  | 2.10 | 0.62 |  |  |  |  |  |  |  |
| 17 | 5000 | 5000 |  |  | 537.86 |  | 84.40 | 7.89 | 43.06 | 3.56 | 2.08 | 0.01 | 0.06 |  |  |
| 18 | 4328.71 | 5000 |  | 5000 |  |  | 2468.05 |  | 5000 |  | 3637.23 |  |  | 114.93 | 12.95 |
| 19 | 5000 |  |  | 344.33 |  |  |  |  |  |  |  |  |  |  |  |
| 20 |  | 5000<sup>a</sup> |  |  | 5000 | 97.79 |  | 344.12 | 5000 |  |  |  | 4664.69 |  |  |
| 21 | 1727.71 | 2393.09 |  | 1354.45 |  |  | 2.51 | 1.61 | 1.05 | 2.59 | 1.03 | 0.01 |  |  |  |
| 22 | 5000 |  | 5000 |  |  | 7.42 | 8.09 | 1.97 | 4.39 | 0.71 | 2.78 | 0.67 | 1.72 | 0.08 | 0.24 |
| 23 |  | 88.50 | 2.89 | 0.09 |  |  |  |  |  |  |  |  |  |  |  |
| 24 | 5000 | 5000 |  |  | 223.72 |  | 2.37 | 8.01 | 5.99 | 41.02 |  | 1.23 | 0.01 |  |  |
| 25 | 5000 |  | 267.69 |  | 38.25 |  |  |  | 84.65 | 301 |  |  | 38.23 | 2.40 | 8.13 |
| 26 | 1305.00 | 5000 |  |  | 1021.42 |  |  |  | 19.95 | 1.89 | 0.57 | 0.24 |  |  |  |
| 27 | 5000 |  |  | 1767.49 | 274.52 | 245.85 | 147.76 | 111.48 |  |  | 3.44 | 6.81 | 1.92 | 1.15 | 0.56 |
| 28 |  | PCR<br>Ct 16.6 | 5000 |  |  | 168.50 |  |  | 86.65 | 1.39 | 14.97 | 6.59 | 496.04 |  |  |
| 29 | 1635.16 |  | 115.32 |  | 1463.78 |  | 41.1 |  |  |  |  |  |  |  |  |
| 30 | 5000 |  | 1125.20 |  |  |  |  |  |  |  |  |  |  |  |  |
| 31 | 5000 | 5000 |  | 5000 |  | 1330.12 |  |  |  |  |  |  |  |  |  |
| 32 | 5000 | 5000 |  | 1292.38 |  |  | 63.29 |  | 9.86 | 11.31 | 1.56 | 0.01 |  |  |  |

Ct cycle threshold, EOT end of treatment, PCR polymerase chain reaction
<sup>a</sup>PCR positive at day 0
Patients received ensitrelvir on days 0–4. Shading represents days the patient was hospitalized

Viral rebound after end of ensitrelvir treatment was observed in four (12.5%) patients: two (Patient 9 and Patient 14) had neither rebound of clinical symptoms, nor additional antiviral treatment; one (Patient 25) had no rebound of clinical symptoms but received remdesivir from day 8 to day 13; and one (Patient 28) experienced transient increase of body temperature ≧ 37.5 degree Celsius at the time of viral rebound and received remdesivir treatment from day 11 to day 15, with viral clearance observed on day 14 and day 15. No disease progression was observed in these four cases. In a subgroup analysis of 21 patients who switched to ensitrelvir following failure of ≧ 3 days remdesivir treatment or remdesivir plus casirivimab/imdevimab, 13/21 (61.9%) achieved viral clearance by day 5. Patient 18 did not achieve viral clearance and received remdesivir between day 7 and day 11, as well as meropenem for aspirational pneumonia with onset after completion of ensitrelvir treatment.

### Secondary clinical outcomes and exploratory endpoints

All 32 patients achieved clinical improvement as assessed by the investigator at the end of ensitrelvir therapy (day 4 following administration of ensitrelvir). Six (18.8%) patients (Patient 2, 4, 17, 18, 27 and 28) experienced a transient increase of body temperature to ≧37.5 degree Celsius after end of ensitrelvir treatment. No patients were admited to the ICU and no patients died up to 14 days following administration of ensitrelvir. In addition, no patients progressed to severe disease over the course of the study. Sixteen patients were discharged from hospital before day 14, and fifteen were discharged after day 14 (range: day 14–77). One patient (Patient 24) died on day 59; this death was considered unrelated to COVID-19 and due to an underlying comorbidity of ANCA-associated vasculitis.

### Safety

No ensitrelvir treatment-related AEs were observed, and ensitrelvir treatment was not discontinued for any patient.

## DISCUSSION

Here we report the first case series for hospitalized patients treated with ensitrelvir for SARS-CoV-2 infection following emergency approval in November 2022. Whilst ensitrelvir has demonstrated rapid alleviation of symptoms in its clinical studies[39, 40, 47], the efficacy of ensitrelvir in patients hospitalized with COVID-19 is unclear. Also, the current Japanese guideline on COVID-19 treatment recommends ensitrelvir for the treatment of mild/moderate I disease only, regardless of the presence of any risk factors for severe disease[42]. This case series represents clinical efficacy of ensitrelvir treatment in hospitalized patients with SARS-CoV-2 in a real-world setting, and has demonstrated successful ensitrelvir use in this population, including patients with moderate II disease severity. This study included a high-risk patient population who had severe comorbidities and an average age of 73.5 years. Despite this, all patients achieved clinical improvement as assessed by the investigator at day 4 and no cases of ICU admission or death occurred over 14 days, indicating that ensitrelvir successfully prevented progression to severe disease in this high-risk group.

In this case series, ensitrelvir also demonstrated antiviral activity; viral clearance (defined in the methods section) was achieved by day 5 in 18/32 (56.3%) patients. Viral persistence is an unmet need for patients with COVID-19, particularly for immunocompromised patients[25, 26] and patients with comorbidities[28, 29] for whom COVID-19 infection is associated with negative outcomes[21–23, 48, 49]. Prolonged SARS-CoV-2 infection has been linked with rapid viral evolution[50, 51], and it has also been hypothesized that viral persistence may increase the likelihood of long COVID[52–54]. Furthermore, reductions in viral titer and load following antiviral therapy have been linked to reduction in mortality rates and improved symptom resolution[34, 55, 56] and may decrease the potential for SARS-CoV-2 transmission[55]. Therefore, it is important to reduce the viral load of SARS-CoV-2, both in the short and long term.

Whilst ensitrelvir in hospitalized patients is being investigated in the ongoing STRIVE study[41], the results of this case series align with reported clinical trial data for ensitrelvir in non-hospitalized patients. In the randomized, placebo-controlled, phase 3 SCORPIO-SR study, reduction from baseline in SARS-CoV-2 viral RNA on day 4 was significantly greater with ensitrelvir compared with placebo (least-squares mean [log_10_ TCID_50_/mL]: −2.48 versus −1.01, *p* < .001)[39]. Information on viral RNA reduction in hospitalized patients for other oral antivirals is limited, but a recent phase 3 study of nirmatrelvir/ritonavir in hospitalized adult patients with severe comorbidities reported no significant difference in duration of SARS–CoV-2 RNA clearance compared with placebo[57]. Not all patients achieved viral clearance following ensitrelvir treatment in this case series, but all patients improved clinically as determined by the investigator at day 4.

Of note, most patients (27/32) in this case series were treated with ensitrelvir after failure of remdesivir treatment (remdesivir alone or remdesivir plus casirivimab/imdevimab). Of these patients, 21 switched to ensitrelvir following failure of ≥ 3 days remdesivir treatment. All patients in this subgroup achieved investigator-judged clinical improvement at day 4, suggesting ensitrelvir is effective even when received ≥ 3 days after onset of infection, an outcome that has not yet been demonstrated in clinical trials[58]. These findings may mean that ensitrelvir could provide clinical benefit to patients even when it is administered at a delayed phase of the disease with persistent viral infection.

Currently available treatments, remdesivir and nirmatrelvir are not recommended for patients with eGFR <30mL/min. In this case series, seven patients received hemodialysis alongside ensitrelvir treatment, all of whom clinically improved, with no adverse event. Of the seven who received hemodialysis, three reported viral clearance by day 5 and five were discharged from hospital by day 14. This suggests that ensitrelvir may be a viable treatment option for these patients.

This retrospective chart review has several limitations. It was a single center study, with a relatively small patient population and, given the nature of the study, there was no control group. As such, it is difficult to draw conclusions on the comparative efficacy and safety of ensitrelvir from this study alone. Secondly, due to the retrospective nature of the study, the evaluation and timings for each outcome do not necessarily match between patients. Not all patients had an antigen quantitative measurement taken after end of ensitrelvir treatment, nor did patients receive them on the same days. Additionally, although these were hospitalized patients, and so represent those impacted heavily by infection, most patients had mild disease severity on commencement of ensitrelvir. There were less than 10 patients in each of the moderate I and moderate II disease groups, limiting the conclusions we can draw from these populations. Finally, there were no pre-specified or standardized criteria for: patients switching to ensitrelvir from remdesivir; the evaluation of clinical outcomes following ensitrelvir treatment; or discharge from hospital following ensitrelvir treatment. All decisions were based on the clinical judgement of the investigator.

The main strength of this study is that it reflects real-world use of ensitrelvir and outcomes in patients with a wide range of comorbidities. The study was also conducted after emergency approval of ensitrelvir in November 2022, during the Omicron era, representing the efficacy of ensitrelvir against this persisting variant.

Whilst ensitrelvir demonstrated efficacy in the majority of patients, there were cases where ensitrelvir failed to achieve viral clearance and cases of viral rebound and/or transient increase of body temperature. Three patients received post-antiviral treatment (remdesivir). There are several reports about rebound of SARS-CoV-2 and symptoms with nirmatrelvir–ritonavir and molnupiravir in hospitalized patients with COVID-19 [59], as well as non-hospitalized patients[60]. In this case series, only one patient developed both viral rebound and a transient increase in body temperature, but did not develop any other symptoms at the time of viral rebound. Overall, in this case series, viral rebound was not correlated with transient increase of body temperature, and did not lead to serious clinical outcomes, which is consistent with previous reports with other SARS-CoV-2 antivirals [59, 61]. Further investigation of antiviral and clinical efficacy of ensitrelvir in hospitalized patients is warranted.

## CONCLUSIONS

Ensitrelvir showed potent antiviral activity and was associated with positive clinical outcomes in a generally high-risk, hospitalized, elderly patient population with mild–moderate COVID-19 infection, including in those with a variety of comorbidities. In the majority of cases, ensitrelvir demonstrated potent antiviral activity despite previous remdesivir treatment failure and clinical improvement was reported in all patients. Of note, no patients progressed to severe or critical diseases of COVID-19 in this generally high-risk population. This case series adds to the growing clinical information supporting the efficacy and safety of ensitrelvir for the treatment of patients infected with SARS-CoV-2, including those hospitalized with a range of high-risk factors, even when treatment was delayed beyond 72 hours from disease onset.

## Data Availability

All associated data are published within the main body of the manuscript and supplementary materials.

## ACKNOWLEDGEMENTS

The authors would like to thank the patients and their families. Rinku General Medical Center used SIMPRESEARCH^®^ as a data tabulation tool, and we would like to thank the SIMPRESEARCH^®^ developer, 4DIN Ltd. The authors would also like to thank Tsukasa Horiyama for their contribution to manuscript development.

## Funding

The implementation of this research was funded by Shionogi & Co., Ltd., Osaka, Japan. Shionogi & Co., Ltd. Also took in charge all costs associated with the development and publication of this manuscript, including the journal’s Rapid Service Fee.

## Medical Writing and Editorial Assistance

Editorial assistance in the preparation of this manuscript, under the direction of the authors, was provided by Ian McAllister, BSc, of Ashfield MedComms, an Inizio Company, funded by Shionogi & Co., Ltd., and complied with Good Publication Practice (GPP) guidelines (DeTora LM, et al. Ann Intern Med 2022;175:1298–1304).

## Author Contributions

Masaya Yamato was responsible for research planning, study conception and design, acquisition of data, quality assurance, interpretation of results, drafting the article, and revising the article critically for important intellectual content. Masayuki Seki and Tomoki Mizuno were responsible for acquisition of data, quality assurance, drafting the article, and revising the article critically for important intellectual content. Masahiro Kinoshita, Shogo Miyazawa and Tomoki Sonoyama were responsible for research planning, interpretation of results, drafting the article, and revising the article critically for important intellectual content. All authors gave final approval before submission.

## Disclosures

Masaya Yamato has received lecture fees from, and serves as an advisor for, Shionogi & Co., Ltd.. Masahiro Kinoshita, Shogo Miyazawa and Takuhiro Sonoyama are employees of Shionogi & Co., Ltd.. Masayuki Seki and Tomoki Mizuno have nothing to disclose. This study was supported by Shionogi & Co., Ltd. In the form of all costs associated with the development and publication of this manuscript.

## Compliance with Ethics Guidelines

The study was conducted in accordance with the Declaration of Helsinki, and approved by the Rinku General Medical Center Clinical Research Ethics Commitee (2-23 Rinku Oraikita, Izumisano City 598-0048; protocol ID: 2023FY-I-001; date of approval 18 May 2023). Patient consent was acquired using an opt-out procedure and no data were included that would allow identification of individual patients. This study was registered in UMIN Clinical Trials Registry (study ID: UMIN000051300).

---

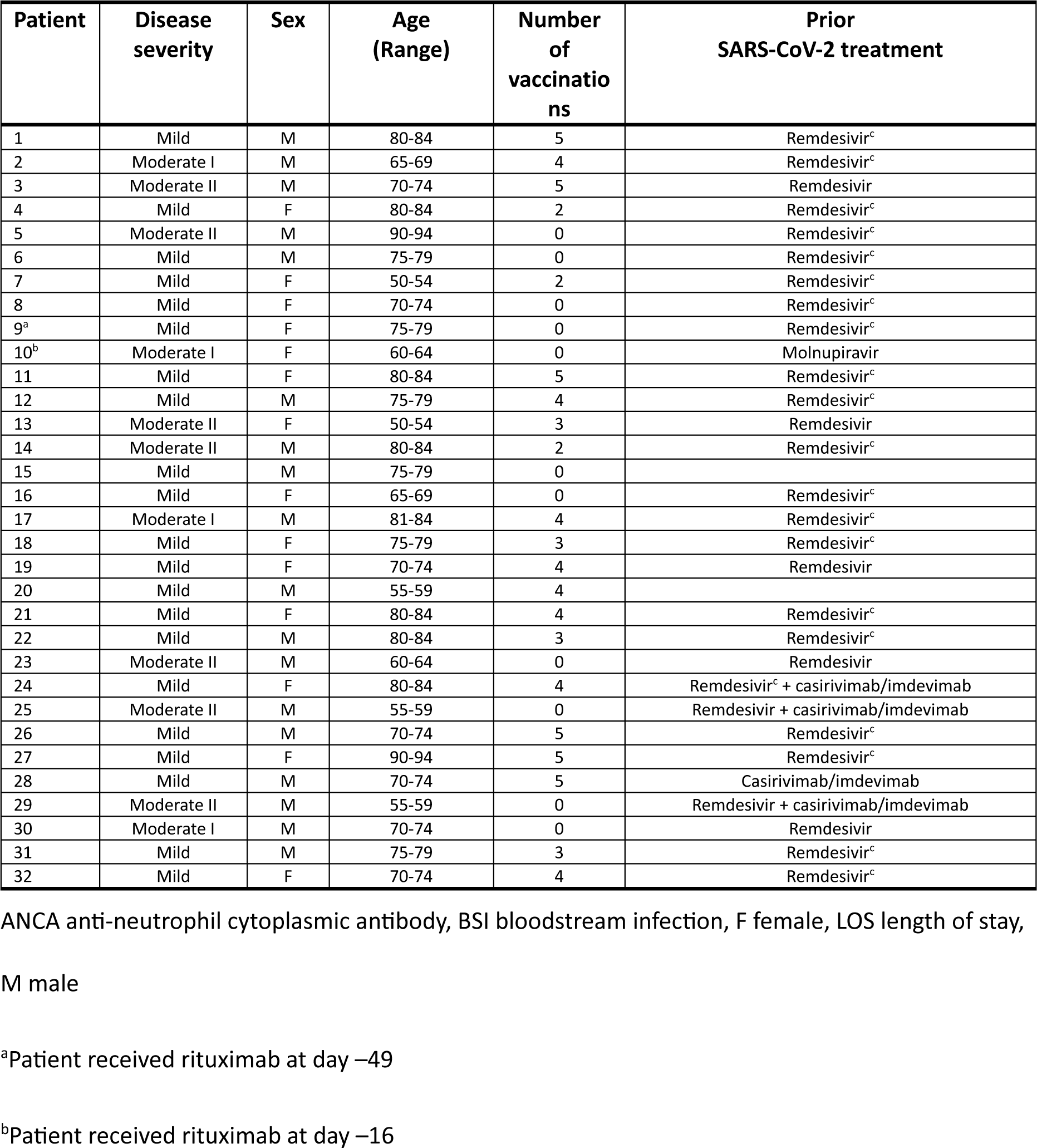
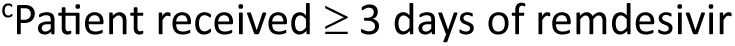

| Patient | Disease severity | Sex | Age (Range) | Number of vaccinations | Prior SARS-CoV-2 treatment |
| --- | --- | --- | --- | --- | --- |
| 1 | Mild | M | 80-84 | 5 | Remdesivir<sup>c</sup> |
| 2 | Moderate I | M | 65-69 | 4 | Remdesivir<sup>c</sup> |
| 3 | Moderate II | M | 70-74 | 5 | Remdesivir |
| 4 | Mild | F | 80-84 | 2 | Remdesivir<sup>c</sup> |
| 5 | Moderate II | M | 90-94 | 0 | Remdesivir<sup>c</sup> |
| 6 | Mild | M | 75-79 | 0 | Remdesivir<sup>c</sup> |
| 7 | Mild | F | 50-54 | 2 | Remdesivir<sup>c</sup> |
| 8 | Mild | F | 70-74 | 0 | Remdesivir<sup>c</sup> |
| 9<sup>a</sup> | Mild | F | 75-79 | 0 | Remdesivir<sup>c</sup> |
| 10<sup>b</sup> | Moderate I | F | 60-64 | 0 | Molnupiravir |
| 11 | Mild | F | 80-84 | 5 | Remdesivir<sup>c</sup> |
| 12 | Mild | M | 75-79 | 4 | Remdesivir<sup>c</sup> |
| 13 | Moderate II | F | 50-54 | 3 | Remdesivir |
| 14 | Moderate II | M | 80-84 | 2 | Remdesivir<sup>c</sup> |
| 15 | Mild | M | 75-79 | 0 |  |
| 16 | Mild | F | 65-69 | 0 | Remdesivir<sup>c</sup> |
| 17 | Moderate I | M | 81-84 | 4 | Remdesivir<sup>c</sup> |
| 18 | Mild | F | 75-79 | 3 | Remdesivir<sup>c</sup> |
| 19 | Mild | F | 70-74 | 4 | Remdesivir |
| 20 | Mild | M | 55-59 | 4 |  |
| 21 | Mild | F | 80-84 | 4 | Remdesivir<sup>c</sup> |
| 22 | Mild | M | 80-84 | 3 | Remdesivir<sup>c</sup> |
| 23 | Moderate II | M | 60-64 | 0 | Remdesivir |
| 24 | Mild | F | 80-84 | 4 | Remdesivir<sup>c</sup> + casirivimab/imdevimab |
| 25 | Moderate II | M | 55-59 | 0 | Remdesivir + casirivimab/imdevimab |
| 26 | Mild | M | 70-74 | 5 | Remdesivir<sup>c</sup> |
| 27 | Mild | F | 90-94 | 5 | Remdesivir<sup>c</sup> |
| 28 | Mild | M | 70-74 | 5 | Casirivimab/imdevimab |
| 29 | Moderate II | M | 55-59 | 0 | Remdesivir + casirivimab/imdevimab |
| 30 | Moderate I | M | 70-74 | 0 | Remdesivir |
| 31 | Mild | M | 75-79 | 3 | Remdesivir<sup>c</sup> |
| 32 | Mild | F | 70-74 | 4 | Remdesivir<sup>c</sup> |

ANCA anti-neutrophil cytoplasmic antibody, BSI bloodstream infection, F female, LOS length of stay,
M male
<sup>a</sup>Patient received rituximab at day –49
<sup>b</sup>Patient received rituximab at day –16
<sup>c</sup>Patient received $\geq 3$ days of remdesivir

## Notes

### Author Declarations

The study was conducted in accordance with the Declaration of Helsinki, and approved by the Rinku General Medical Center Clinical Research Ethics Committee (2-23 Rinku Oraikita, Izumisano City 598-0048; protocol ID: 2023FY-I-001; date of approval 18 May 2023). Patient consent was acquired using an opt-out procedure and no data were included that would allow identification of individual patients. This study was registered in UMIN Clinical Trials Registry (study ID: UMIN000051300).

